# The epidemiology of long COVID in US adults two years after the start of the US SARS-CoV-2 pandemic

**DOI:** 10.1101/2022.09.12.22279862

**Authors:** McKaylee M Robertson, Saba A Qasmieh, Sarah G Kulkarni, Chloe A Teasdale, Heidi Jones, Margaret McNairy, Luisa N. Borrell, Denis Nash

## Abstract

**Objectives:** To characterize prevalence and impact of long COVID.

**Methods:** We conducted a population-representative survey, June 30-July 2, 2022, of a random sample of 3,042 United States adults. Using questions developed by the United Kingdom’s Office of National Statistics, we estimated the prevalence by sociodemographics, adjusting for gender and age.

**Results:** An estimated 7.3% (95% CI: 6.1-8.5%) of all respondents reported long COVID, approximately 18,533,864 adults. One-quarter (25.3% [18.2-32.4%]) of respondents with long COVID reported their day-to-day activities were impacted ‘a lot’ and 28.9% had SARS-CoV-2 infection >12 months ago. The prevalence of long COVID was higher among respondents who were female (aPR: 1.84 [1.40-2.42]), had comorbidities (aPR: 1.55 [1.19-2.00]) or were not (versus were) boosted (aPR: 1.67 [1.19-2.34]) or not vaccinated (versus boosted) (aPR: 1.41 (1.05-1.91)).

**Conclusions:** We observed a high burden of long COVID and substantial variability in prevalence of SARS-CoV-2. Population-based surveys are an important surveillance tool and supplement to ongoing efforts to monitor long COVID.

## INTRODUCTION

Post-COVID conditions (PCC) or “long COVID” consists of new, recurring or ongoing symptoms or clinical findings four or more weeks after SARS-CoV-2 infection, and may potentially affect millions of the 58% of Americans estimated to have been infected with COVID since March of 2020.^1–4^ Estimates of the prevalence of long COVID vary widely based on study design, population, and case definition.^5^ The United States (US) Centers for Disease Control and Prevention estimated that one in five people with *prior* COVID are currently experiencing long COVID as of June 2022^6^. In the US, assessments of long COVID prevalence have largely been derived from sources that are not population-representative: cross-sectional convenience samples or electronic health record and cohort analyses.^2,3,6–8^ Routinely updated, population-representative estimates of long COVID prevalence are available in the United Kingdom (UK)^9^ where the Office of National Statistics (ONS) assesses long-COVID symptoms approximately every four weeks. Over the 4-week period ending June 4, 2022, an estimated 2 million (2.8%) of the total UK population aged 2 years or older had long COVID.^9^

Little is known about risk factors for long COVID and the impact of long COVID on activities of daily living. Knowledge gaps include who is most vulnerable and the extent to which vaccines and boosters may be protective. In data from the US and Europe, long COVID prevalence following SARS-CoV-2 infection appears to be consistently higher among women and among people with multiple chronic conditions.^5,10–13^ However, beyond female sex and presence of chronic conditions, data on risk factors vary, likely due to differences in a) study design, often lacking COVID-free or other control/comparison groups, b) population, often conducted among hospitalized or healthcare seeking populations, and/or c) case definition of long COVID.^10–12,14,15^ Given that SARS-CoV-2 vaccines, especially without boosters, are increasingly less effective at providing sterilizing immunity against omicron variants^16–20^ and that half of the population is estimated to have had COVID at least once^4^, understanding the prevalence of long COVID by vaccination status is critical. Limited data suggest a modest reduction in long COVID risk among vaccinated compared with unvaccinated or COVID-naive people and among unvaccinated people who went on to receive a vaccination versus people who did not.^21,22^

Our objective was to characterize the prevalence and risk factors of long COVID and the impact of long COVID on daily living in a population representative sample of US adults (aged 18+ years).

## METHODS

### Study sample

We conducted a probability-based, cross-sectional survey, in English and Spanish, between June 30 and July 2, 2022, of 3,042 adults living in the United States (US). Two stratified proportionate random population-based samples of potential participants were drawn from a sampling frame of 105,469,157 mobile numbers and from a frame 60,126,257 landlines. To help ensure representativeness, a sample from a National opt-in Online Panel was also drawn. In the final sample, 62% of respondents were recruited from landline, 32% from mobile and 6% from the online opt-in panel. To create weights representative of the US-population 18 years or older, we used an iterative weighting method, raking, to marginal proportions of race, ethnicity, age, self-identified sex, and education by US region based on the 2020 US census^23^. Our overall response rate across all modalities was 7.2%, which is comparable with the Household Pulse Survey (HPS), a national online survey sampling households from the Census Master Address File with an email or cell phone number. Further details on the survey design, sampling and weighting are provided in Appendix 1.

### Prevalence of ever having SARS-CoV-2 as of July 2022

We assessed current and previous SARS-CoV-2 infection status. Respondents with a current SARS-CoV-2 infection during the 14-days prior to the survey were identified based on the following mutually exclusive, hierarchical case classification: 1) self-report of one or more positive tests with a health care or testing provider; or 2) self-report of a positive test result exclusively based on at-home rapid tests (i.e. those that were not followed up with confirmatory diagnostic testing with a provider); or 3) self-report of COVID-like symptoms AND a known epidemiologic link (close contact) to one or more laboratory confirmed or probable (symptomatic) SARS-CoV-2 case(s)^24^ in a respondent who reported never testing or only testing negative during the study period. To assess previous infections we asked all participants “Prior to June 15th 2022, did you ever have COVID-19 infection, either diagnosed by a healthcare or testing provider, or based on a positive at-home rapid test” and the most recent time they had COVID. The prevalence of ever having COVID as of July 2022 was estimated as either meeting the case definition for a current SARS-CoV-2 infection or responding affirmatively to having SARS-CoV-2 infection prior to July 15th, 2022. The full survey questionnaire is in Appendix 2.

### Long COVID Prevalence Estimation

We used a question asked routinely by the ONS in the UK to define and assess the burden of long COVID.^25^ Participants who reported any history of prior COVID were also asked “Would you describe yourself as having ‘long COVID’, that is you experienced symptoms such as fatigue, difficulty concentrating, shortness of breath more than 4 weeks after you first had COVID-19 that are not explained by something else?” The prevalence of long COVID was estimated as the proportion responding affirmatively. Respondents whose most recent SARS-CoV-2 infection was within the past month were classified as not having long COVID, to avoid conflation of current symptoms with recent acute illness and to align with the UK definition of long COVID, which was assessed more than four weeks from diagnosis. We examined prevalence among all respondents, with or without prior COVID, to compare with the UK ONS estimate.

### Reduced ability to carry out daily activities with long COVID

We included a question used by the UK ONS to assess the impact of long COVID on daily living. Participants who reported that they had long COVID were asked “Does this reduce your ability to carry-out day-to-day activities compared with the time before you had COVID-19?” Response choices were on a likert 3-point scale of: “Yes, a lot; Yes, a little;” or “Not at all”.

### Predictors of long COVID

In addition to sociodemographic factors, we assessed other potential predictors of long COVID status. To assess the presence or absence of comorbidities, we asked “do you have any of the following conditions that could increase the severity of COVID-19: cancer, diabetes, obesity, COPD or lung disease, liver disease, heart disease, high blood pressure, a recent organ transplant, or an immunodeficiency)?” The variable was dichotomized as Yes versus No/Do not know. We collected information on whether, at the time of interview, participants were fully vaccinated or boosted at least once. Respondents were classified as boosted, fully vaccinated and not boosted, or not vaccinated/partially vaccinated.

### Statistical Analysis

We estimated the prevalence of having ever had COVID as of July 2022, having long COVID and its impact to daily living by socio-demographic characteristics, geographic region, current vaccination status, timing of most recent infection, and presence of comorbidities. Survey weights were applied to generate estimates of the proportion with COVID or long COVID along with 95% confidence interval (95%CI). We used direct standardization to calculate age and sex-adjusted prevalence estimates using the US 2020 census.^23^

We used log-binomial models to obtain prevalence ratios (PR) and 95% confidence intervals by sociodemographic characteristics or region, adjusting for age and gender (aPR). As a sensitivity analysis of the age and gender-adjusted models, and given these models were predictive not explanatory, we also used backward selection, keeping all significant predictors. For impact of long COVID on daily living, among the subgroup with long COVID, we compared “A lot” of reduced ability (versus a little and none). Due to smaller sample sizes some sociodemographic factors were collapsed into larger categories (e.g., ages 35-49). We used gender for adjustment since our survey captured only gender. For standardization, we had to assume that reported gender was reported sex.

The study protocol was approved by the Institutional Review Board at the City University of New York (CUNY IRB 2022-0407).

## RESULTS

### Prevalence of and risk factors for Long COVID in the general adult population

An estimated 7.3% (95% CI 6.1-8.5%) of all respondents reported currently having long COVID as of July 2, 2022, which corresponds to approximately 18,533,864 US adults (Table 1). The standardized prevalence of long COVID was highest among respondents who were aged 25-34 (10.0% [6.8-14.6%]), 35-44 (9.0% [6.1-13.1%]), female (9.4% [7.7-11.6%]), White non Hispanic (NH) (8.7% [7.1-10.7%]), had a household income of $20,001-60,000 (8.8% [6.6-11.6%]) or 60,001-100,000 (8.5% [6.1-11.7%]), were employed (8.9% (7.2-11.0%), or reported having comorbidities (10.3% [7.8-13.4%]) (Table 1). The standardized prevalence of long COVID was lowest among respondents who were youngest (18-24 years: 4.4% [1.8-10.0%]), oldest (65+ years: 4.3% (3.0-5.8%]), male (5.0% [3.8-6.5%]), Black NH (5.2% [2.9-9.3%]), Hispanic (5.7% [2.9-11.0%]), Asian/Pacific Islander (API) NH (1.4% [0.3-5.9%]), uninsured (4.3% [2.7-6.8%]) or currently vaccinated and boosted (5.8% [4.4-7.7%]).

**Table 1.** Prevalence and characteristics of US adults with COVID more than one month ago and long COVID, July 2022 - Among Wtd N = 3042 Respondents

|  |  | Prevalence of Long COVID Among All Respondent (with or without COVID) |  |  |  |  |  |
| --- | --- | --- | --- | --- | --- | --- | --- |
|  | Total | Long COVID | Crude Prevalence of Long COVID % (95% CI) | Age and sex direct-standardized prevalence of long COVID* % (95% CI) | Crude prevalence ratio (PR) PR (95% CI) | Adjusted prevalence ratio (aPR)** aPR (95% CI) | Estimated Number with Long COVID |
|  | Weighted N (%) | Weighted N (%) |  |  |  |  |  |
| **Total** | 3,042 (100.0) | 222 (100.0) | 7.3 (6.1, 8.5) |  |  |  | 18,533,864 |
| **Age** |  |  |  |  |  |  |  |
| 18-24 | 365 (12.0) | 17 (7.8) | 4.8 (0.9, 8.6) | 4.4 (1.8 - 10.0) | **0.45 (0.27, 0.76)** | **0.50 (0.30, 0.84)** | 1,445,641 |
| 25-34 | 547 (18.0) | 58 (26.1) | 10.6 (6.6, 14.6) | 10.0 (6.8 - 14.6) | Ref | Ref | 4,837,339 |
| 35-44 | 495 (16.3) | 43 (19.1) | 8.6 (5.3, 11.9) | 9.0 (6.1 - 13.1) | 0.81 (0.56, 1.18) | 0.87 (0.60, 1.26) | 3,539,968 |
| 45-54 | 498 (16.4) | 37 (16.5) | 7.4 (4.6, 10.2) | 7.4 (5.1 - 10.7) | 0.70 (0.47, 1.03) | 0.72 (0.49, 1.07) | 3,058,088 |
| 55-64 | 508 (16.7) | 41 (18.2) | 8.0 (5.5, 10.5) | 8.3 (6.0 - 11.2) | 0.75 (0.51, 1.10) | 0.80 (0.55, 1.18) | 3,373,163 |
| 65+ | 629 (20.7) | 27 (12.2) | 4.3 (3.0, 5.6) | 4.2 (3.0 - 5.8) | **0.41 (0.26, 0.63)** | **0.43 (0.27, 0.66)** | 2,261,131 |
| **Gender** |  |  |  |  |  |  |  |
| Male | 1,443 (47.4) | 72 (32.4) | 5.0 (3.6, 6.3) | 5.0 (3.8 - 6.5) | Ref | Ref | 6,004,972 |
| Female | 1,516 (49.8) | 144 (64.8) | 9.5 (7.5, 11.5) | 9.4 (7.7 - 11.6) | **1.90 (1.45, 2.50)** | **1.84 (1.40, 2.42)** | 12,009,944 |
| Non-binary*** | 82 (2.7) | 6 (2.8) | 7.4 (0.0, 17.2) | 6.3 (2.1 - 17.6) | 1.49 (0.67, 3.30) | 1.45 (0.66, 3.21) | 518,948 |
| **Race/Ethnicity** |  |  |  |  |  |  |  |
| Black NH | 350 (11.5) | 19 (8.4) | 5.3 (2.1, 8.5) | 5.2 (2.9 - 9.3) | **0.64 (0.40, 1.03)** | **0.60 (0.38, 0.96)** | 1,556,845 |
| White NH | 1,794 (59.0) | 148 (66.8) | 8.3 (6.8, 9.8) | 8.7 (7.1 - 10.7) | Ref | Ref | 12,380,621 |
| Hispanic | 460 (15.1) | 22 (9.7) | 4.7 (1.3, 8.2) | 5.7 (2.9 - 11.0) | **0.57 (0.37, 0.88)** | **0.57 (0.37, 0.89)** | 1,797,785 |
| Asian/Pacific Islander | 171 (5.6) | 7 (3.2) | 4.2 (0.0, 9.4) | 1.4 (0.3 - 5.9) | 0.51 (0.25, 1.06) | **0.44 (0.21, 0.93)** | 593,084 |
| Other | 268 (8.8) | 26 (11.8) | 9.8 (4.4, 15.2) | 11.3 (6.8 - 18.3) | 1.18 (0.80, 1.76) | 1.23 (0.82, 1.86) | 2,186,996 |
| **Education** |  |  |  |  |  |  |  |
| Some college, college graduate or above | 1,859 (61.1) | 138 (61.9) | 7.4 (6.0, 8.8) | 6.6 (4.9 - 8.9) | 1.04 (0.80, 1.34) | 1.08 (0.83, 1.41) | 11,472,462 |
| High school or below | 1,183 (38.9) | 85 (38.1) | 7.2 (4.9, 9.4) | 7.6 (6.2 - 9.2) | Ref | Ref | 7,061,402 |
| **Household income** |  |  |  |  |  |  |  |
| Below 20K | 561 (18.4) | 34 (15.4) | 6.1 (3.2, 9.0) | 5.8 (3.7 - 9.1) | 0.88 (0.55, 1.39) | 0.86 (0.54, 1.38) | 2,854,215 |
| 20,001 - 60,000 | 941 (30.9) | 83 (37.2) | 8.8 (6.3, 11.3) | 8.8 (6.6 - 11.6) | 1.26 (0.85, 1.86) | 1.13 (0.76, 1.68) | 6,894,598 |
| 60,001 - 100,000 | 568 (18.7) | 48 (21.8) | 8.5 (5.8, 11.3) | 8.5 (6.1 - 11.7) | 1.22 (0.80, 1.88) | 1.15 (0.75, 1.76) | 4,040,382 |
| Above 100,000 | 460 (15.1) | 32 (14.4) | 7.0 (4.2, 9.7) | 7.1 (4.7 - 10.4) | Ref | Ref | 2,668,876 |
| Prefer not to answer | 511 (16.8) | 25 (11.1) | 4.8 (2.4, 7.3) | 4.8 (2.9 - 7.8) | 0.69 (0.42, 1.15) | 0.75 (0.45, 1.25) | 2,057,259 |
| **Employed July 2022** |  |  |  |  |  |  |  |
| Yes | 1,472 (48.4) | 123 (55.3) | 8.3 (6.5, 10.2) | 8.9 (7.2 - 11.0) | **1.32 (1.02, 1.70)** | **1.34 (1.02, 1.76)** | 10,249,227 |
| No/DK | 1,570 (51.6) | 99 (44.7) | 6.3 (4.8, 7.9) | 7.0 (5.2 - 9.5) | Ref | Ref | 8,284,637 |
| **Geographic region** |  |  |  |  |  |  |  |
| Northeast | 541 (17.8) | 29 (13.2) | 5.4 (3.4, 7.5) | 5.6 (3.9 - 8.2) | Ref | Ref | 2,446,470 |
| South | 1,153 (37.9) | 92 (41.5) | 8.0 (6.1, 10.0) | 7.8 (6.1 - 9.8) | 1.47 (0.99, 2.20) | 1.40 (0.94, 2.09) | 7,691,554 |
| Midwest | 630 (20.7) | 52 (23.6) | 8.3 (5.5, 11.2) | 7.6 (5.3 - 10.8) | 1.53 (0.99, 2.37) | 1.52 (0.98, 2.35) | 4,373,992 |
| West | 718 (23.6) | 48 (21.6) | 6.7 (3.8, 9.6) | 6.5 (4.3 - 9.9) | 1.23 (0.79, 1.92) | 1.23 (0.79, 1.91) | 4,003,315 |
| **Current vaccination status** |  |  |  |  |  |  |  |
| Boosted at least once | 1,610 (52.9) | 86 (38.7) | 5.3 (4.0, 6.7) | 5.8 (4.4 - 7.7) | Ref | Ref | 7,172,606 |
| Fully vaccinated not boosted | 476 (15.6) | 49 (22.2) | 10.4 (6.6, 14.1) | 9.8 (6.8 - 13.8) | **1.94 (1.39, 2.71)** | **1.67 (1.19, 2.34)** | 4,114,518 |
| Not vaccinated | 957 (31.5) | 87 (39.2) | 9.1 (6.6, 11.6) | 8.5 (6.4 - 11.3) | **1.71 (1.28, 2.27)** | **1.41 (1.05, 1.91)** | 7,265,275 |
| **Timing of Most Recent COVID infection** |  |  |  |  |  |  |  |
| Never | 1438 (47.3) | NA | NA | NA | NA | NA | NA |
| Within the last month | 568 (18.7) | NA | NA | NA | NA | NA | NA |
| 1-3 months ago | 138 (4.5) | 18 (8.2) | 13.3 (6.9, 19.6) | 14.1 (8.9 - 21.0) | **0.50 (0.31, 0.80)** | **0.48 (0.30, 0.78)** | 1,519,777 |
| 3-6 months ago | 266 (8.7) | 77 (34.8) | 29.1 (21.8, 36.4) | 29.0 (22.8 - 36.0) | 1.09 (0.83, 1.45) | 1.07 (0.81, 1.42) | 6,449,785 |
| 6-12 months ago | 219 (7.2) | 60 (27.1) | 27.5 (19.3, 35.6) | 26.7 (19.8 - 34.9) | 1.03 (0.77, 1.40) | 0.96 (0.71, 1.30) | 5,022,677 |
| >12 months ago | 242 (8.0) | 64 (28.9) | 26.6 (19.7, 33.4) | 26.5 (19.7 - 34.5) | Ref | Ref | 5,356,287 |
| Dont know | 171 (5.6) | 2 (0.9) | 1.2 (0.0, 2.8) | 1.5 (0.3 - 8.1) | **0.05 (0.01, 0.18)** | **0.05 (0.01, 0.18)** | 166,805 |
| **Comorbidities** |  |  |  |  |  |  |  |
| Yes | 1,104 (36.3) | 96 (43.3) | 8.7 (6.6, 10.8) | 10.3 (7.8 - 13.4) | **1.34 (1.04, 1.73)** | **1.55 (1.19, 2.00)** | 8,025,163 |
| No | 1,938 (63.7) | 126 (56.7) | 6.5 (5.0, 8.0) | 6.1 (4.8 - 7.6) | Ref | Ref | 10,508,701 |
| **Insured July 2022** |  |  |  |  |  |  |  |
| Yes | 2,398 (78.8) | 190 (85.7) | 7.9 (6.5, 9.4) | 8.0 (6.6 - 9.6 ) | **1.61 (1.11, 2.31)** | **1.93 (1.33, 2.80)** | 15,883,522 |
| No/DNK | 644 (21.2) | 32 (14.3) | 4.9 (2.6, 7.3) | 4.3 (2.7 - 6.8) | Ref | Ref | 2,650,343 |

\*Direct standardized for the age and sex groupings in the 2020 US census
\*\*Model adjusted for sex and age
\*\*\*Estimates are adjusted for age only

In age and gender-adjusted models, there was a higher prevalence of long COVID among respondents who were females (versus males) (aPR: 1.84 [1.40-2.42]), with (versus without) comorbidities (aPR: 1.55 [1.19-2.00]), who were employed (versus unemployed) (aPR: 1.34 [1.02-1.76]), who were insured (versus not or unknown) (aPR: 1.93 [1.33-2.80]), who were not (versus were) boosted (aPR: 1.67 [1.19-2.34]), or who were not vaccinated (versus boosted) (aPR: 1.41 (1.05-1.91)). We observed a lower prevalence of long COVID among respondents who were aged 18-24 (versus 25-34 years)(aPR: 0.50 [0.30-0.84]), aged 65+ (versus 25-34 years) (aPR: 0.43 [0.27-0.66]), were Black NH (versus White NH) (aPR: 0.60 [0.38-0.96]), were Hispanic (versus White NH) (aPR: 0.57 [0.37-0.89]), or were API NH (versus White NH) (aPR: 0.44 [0.21-0.93]). In the backward selection model, which included age, gender, race/ethnicity, current vaccination status, presence of comorbidities or current insurance status, the effect sizes remained the same or increased in strength (further from null) (Supplemental Table 1).

### The impact of long COVID on daily activities

The proportion of respondents with long COVID reporting a reduced ability to carry out daily activities and associations of this reporting with individual characteristics are shown in Table 2. Due to small sample sizes, 95% confidence intervals were wide. One-quarter (25.3% [18.2-32.4%]), an estimated 4.7 million adults, reported their daily activities were impacted ‘a lot’, and reporting of ‘a lot’ of impact was more common among respondents who were aged 50+ (33.6% [23.9-43.2%]), were White NH (33.6% [23.9-43.2%]), had a household income <60K (32.9% [21.4-44.4%]), or were unemployed (36.3% [24.1-48.6%]). In age- and gender-adjusted models, we observed a significantly higher proportion reporting ‘a lot’ of impact on ability of daily activities among respondents with a household income <60K (versus 60K+) (aPR: 2.22 [1.31-3.78]), and a lower proportion reporting ‘a lot’ of reduced ability of daily activities among employed (versus unemployed) respondents (aPR: 0.48 [0.29-0.79]).

**Table 2.**
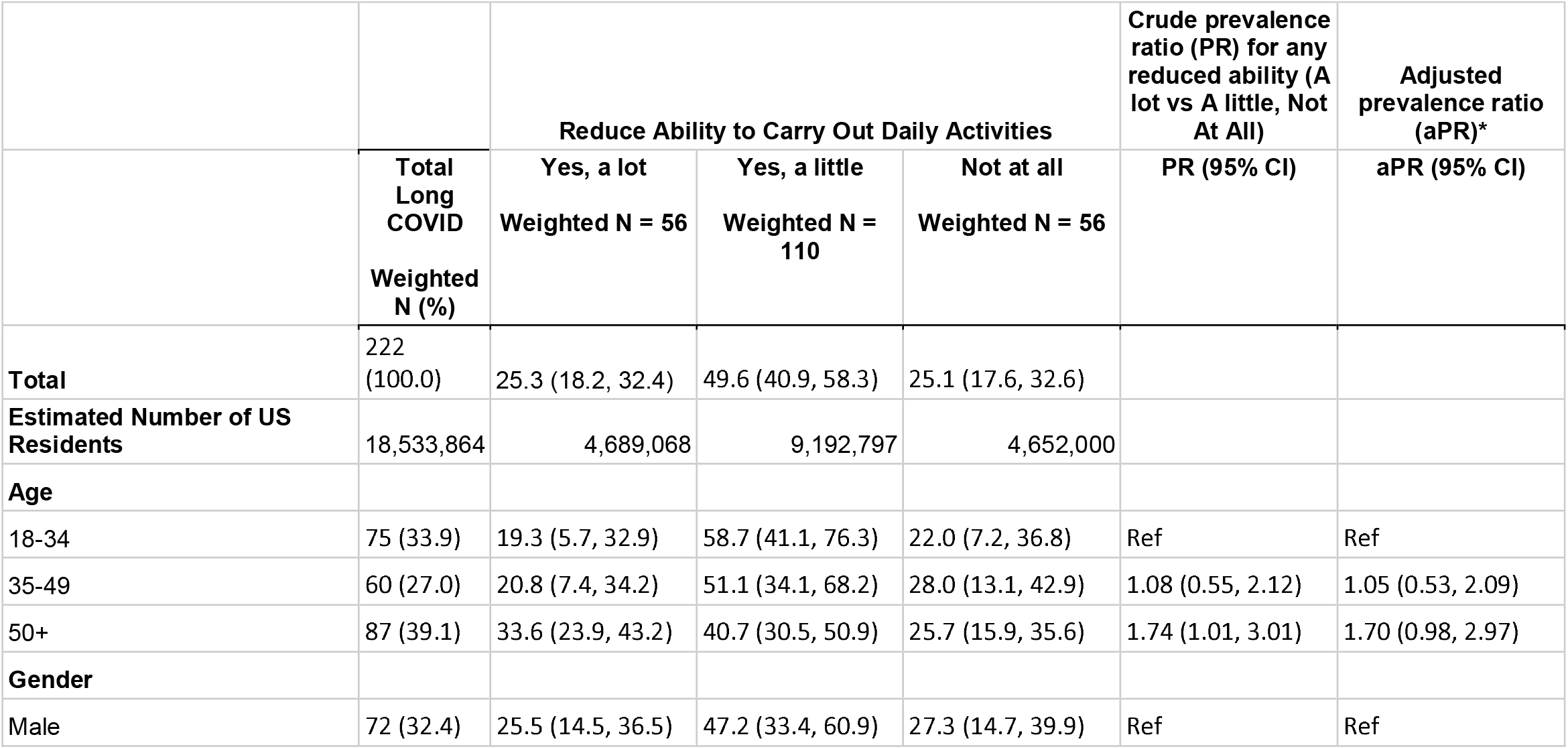
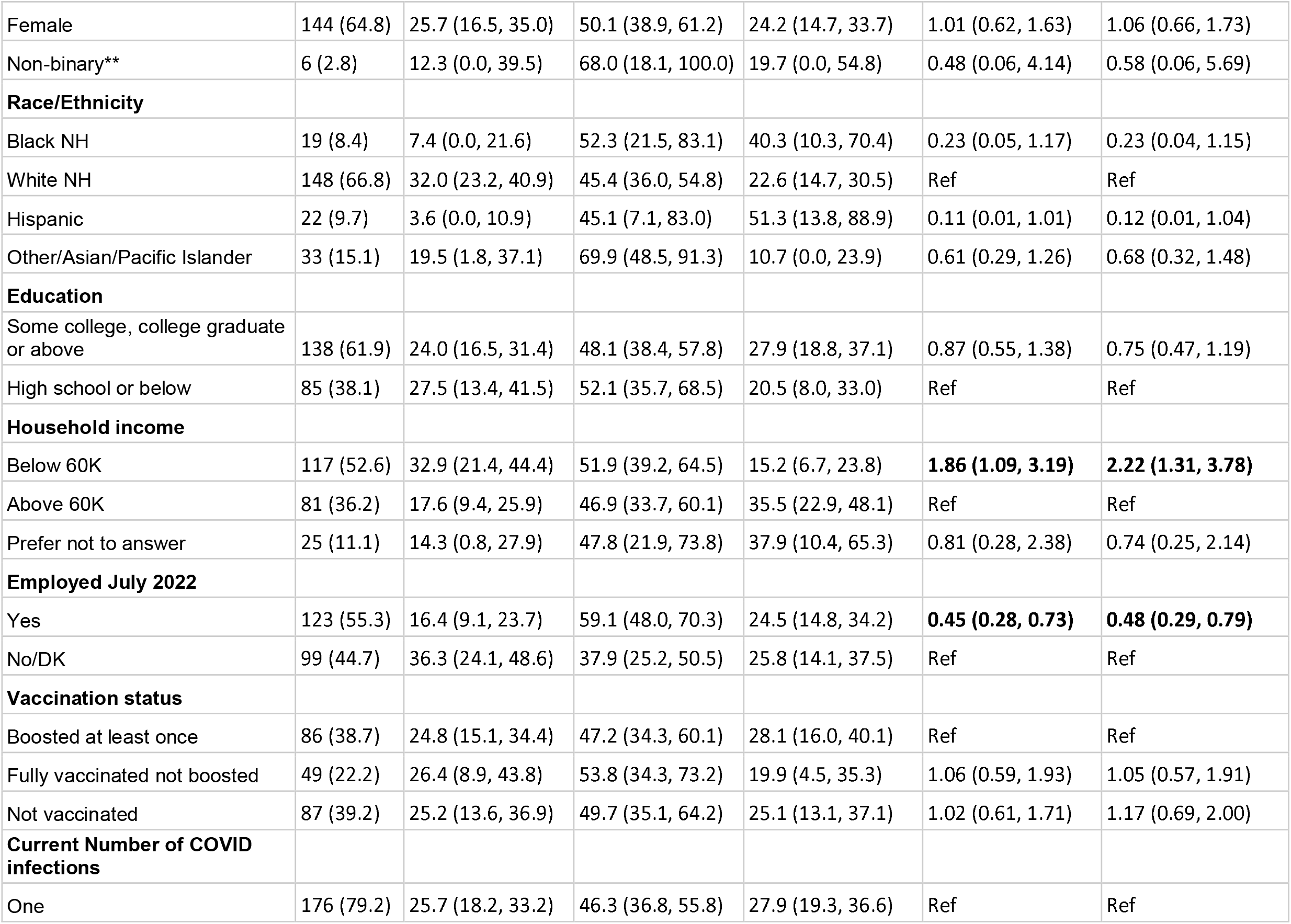
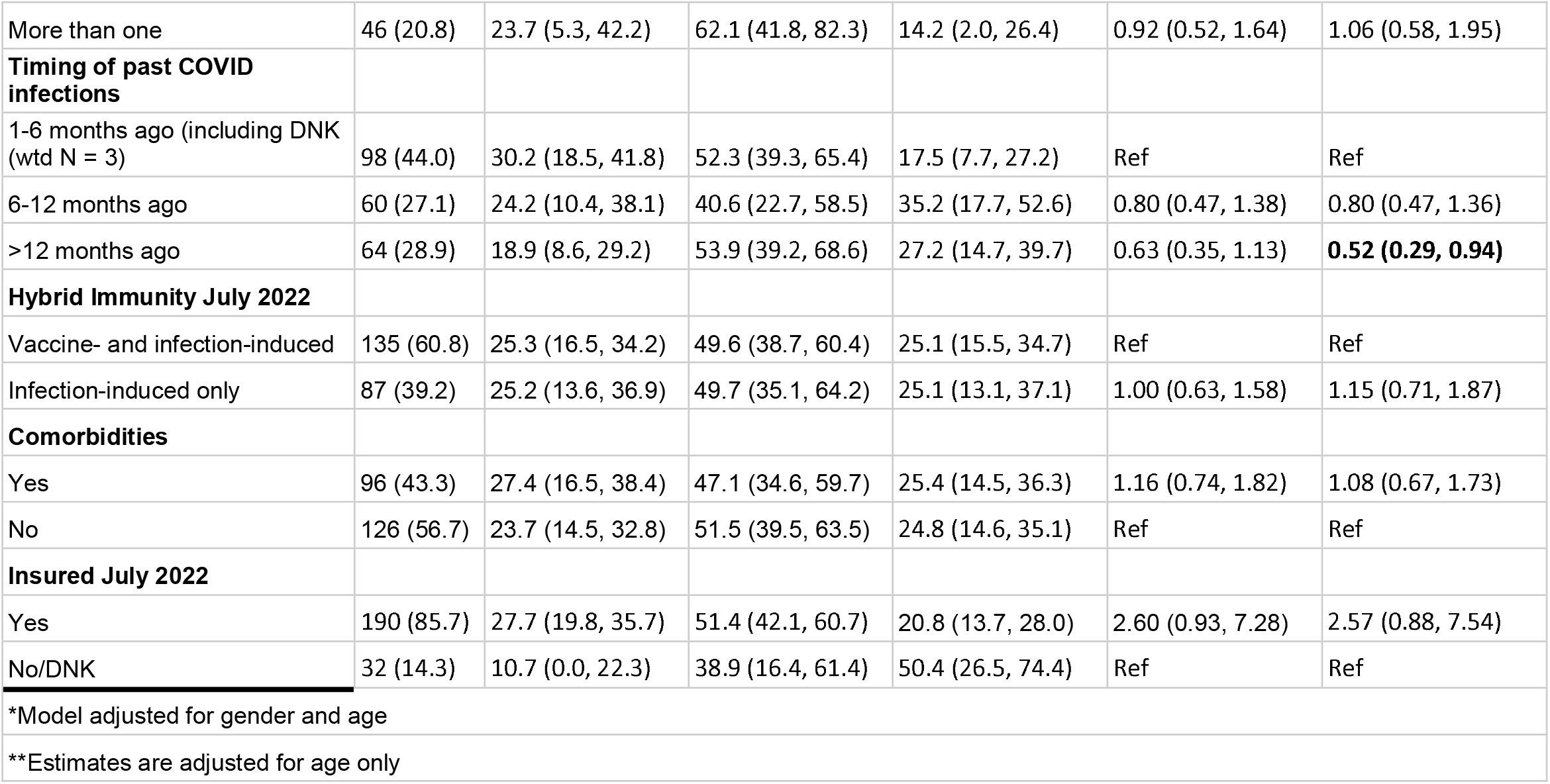
Prevalence and characteristics of US adults with reduced ability to carry out daily activities among those with long COVID, July 2022

Among those with long COVID, less than half (44%) reported their most recent SARS-CoV-2 infection within the past 1-6 months ago, 27% reported within 6-12 months, and 29% >12 months ago. The impact on daily activities decreased modestly with time since the most recent SARS-CoV-2 infection. The proportion reporting ‘a lot’ of impact on daily living was highest among respondents with a SARS-CoV-2 infection within the past 1-6 months (30.2% [18.5-41.8%]) followed by 6-12 months ago (24.2% [10.2-38.1%]; aPR_versus 1-6months_: 0.80 (0.47, 1.36)) and >12 months ago (18.9% [8.6-29.2%]; aPR_versus 1-6months_:0.52 [0.29-0.94]).

### Risk factors for ever having SARS-CoV-2 infection as of July 2022

The prevalence of ever having SARS-CoV-2 infection as of July 2022 was 52.7% (95% CI: 50.2-55.2%) (Table 3). The age- and sex-standardized prevalence of ever having SARS-CoV-2 infection was greatest among the youngest age groups (25-34 years (66.0% [59.0-72.0%]) and decreased with age. The standardized prevalence of ever having SARS-CoV-2 infection was higher among males (56.8% [53.2-60.4%]) than among females (47.8% [44.5-51.1%]) and among those with comorbidities (59.8 [55.8-63.7%]) than those without comorbidities (48.4% [45.4-51.5%]). The standardized prevalence of ever having a SARS-CoV-2 infection was 44.9% (37.9-52.1%) among Black NH respondents, 53.2% (50.4-55.9%) among White NH, 60.7% (52.7-68.1%) among Hispanic, and 34.6% (24.3-46.5%) among API respondents.

**Table 3.** Prevalence and characteristics of US adults with COVID and prevalence of long COVID among US adults with COVID, July 2022

|  |  |  |
| --- | --- | --- |
|  |  | Prevalence of Ever COVID as of July 2022 |

|  | Total | Had COVID<br>as of July<br>2022 | Crude Prevalence<br>of COVID % (95%<br>CI) | Age and sex direct-standardized<br>prevalence of COVID* | Crude<br>prevalence ratio<br>(PR) | Adjusted<br>prevalence ratio<br>(aPR)** |
| --- | --- | --- | --- | --- | --- | --- |
|  | Weighted N<br>(%) | Weighted N<br>(%) |  | % (95% CI) | PR (95% CI) | aPR (95% CI) |
| **Total** | 3,042 (100.0) | 1604 (100.0) | 52.7 (50.2, 55.2) |  |  |  |
| **Age** |  |  |  |  |  |  |
| 18-24 | 365 (12.0) | 235 (14.6) | 64.4 (54.8, 74.0) | 60.8 (51.9 - 69.0) | 0.97 (0.88, 1.07) | 0.98 (0.89, 1.07) |
| 25-34 | 547 (18.0) | 364 (22.7) | 66.5 (60.1, 72.8) | 66.0 (59.0 - 72.4) | Ref | Ref |
| 35-44 | 495 (16.3) | 265 (16.5) | 53.4 (47.0, 59.9) | 51.5 (44.9 - 58.0) | **0.80 (0.73, 0.89)** | **0.81 (0.74, 0.90)** |
| 45-54 | 498 (16.4) | 237 (14.8) | 47.6 (41.3, 53.8) | 47.6 (41.4 - 54.0) | **0.72 (0.64, 0.80)** | **0.72 (0.65, 0.81)** |
| 55-64 | 508 (16.7) | 250 (15.6) | 49.2 (44.4, 54.1) | 49.0 (44.1 - 53.8) | **0.74 (0.67, 0.82)** | **0.74 (0.67, 0.83)** |
| 65+ | 629 (20.7) | 254 (15.8) | 40.3 (37.3, 43.3) | 39.6 (36.6 - 42.7) | **0.61 (0.54, 0.68)** | **0.61 (0.55, 0.68)** |
| **Gender** |  |  |  |  |  |  |
| Male | 1,443 (47.4) | 813 (50.7) | 56.3 (52.5, 60.2) | 56.8 (53.2 - 60.4) | Ref | Ref |
| Female | 1,516 (49.8) | 726 (45.3) | 47.9 (44.6, 51.2) | 47.8 (44.5 - 51.1) | **0.85 (0.79, 0.91)** | **0.84 (0.78, 0.89)** |
| Non-binary*** | 82 (2.7) | 65 (4.0) | 78.5 (66.4, 90.6) | 77.1 (64.1 - 86.4) | 1.39 (1.23, 1.57) | 1.22 (1.09, 1.36) |
| **Race/Ethnicity** |  |  |  |  |  |  |
| Black NH | 350 (11.5) | 169 (10.5) | 48.2 (40.7, 55.6) | 44.9 (37.9 - 52.1) | 0.90 (0.80, 1.02) | **0.81 (0.72, 0.91)** |
| White NH | 1,794 (59.0) | 922 (57.5) | 51.4 (48.9, 53.9) | 53.2 (50.4 - 55.9) | Ref | Ref |
| Hispanic | 460 (15.1) | 307 (19.2) | 66.8 (58.6, 75.0) | 60.7 (52.7 - 68.1) | **1.29 (1.19, 1.40)** | **1.16 (1.07, 1.25)** |
| Asian/Pacific Islander | 171 (5.6) | 75 (4.6) | 43.7 (28.9, 58.5) | 34.6 (24.3 - 46.5) | **0.78 (0.64, 0.95)** | **0.69 (0.57, 0.84)** |
| Other | 268 (8.8) | 131 (8.2) | 48.8 (39.3, 58.3) | 50.7 (40.7 - 60.6) | 0.90 (0.78, 1.04) | 0.88 (0.76, 1.02) |
| **Education** |  |  |  |  |  |  |
| Some college, college graduate or above | 1,859 (61.1) | 942 (58.7) | 50.7 (48.0, 53.3) | 53.4 (49.0 - 57.7) | Ref | Ref |
| High school or below | 1,183 (38.9) | 662 (41.3) | 56.0 (51.2, 60.8) | 51.5 (48.5 - 54.4) | 1.10 (1.03, 1.18) | 1.04 (0.97, 1.11) |
| **Household income** |  |  |  |  |  |  |
| Below 20K | 561 (18.4) | 327 (20.4) | 58.3 (51.1, 65.6) | 54.2 (47.9 - 60.2) | 0.98 (0.88, 1.08) | 0.98 (0.89, 1.08) |
| 20,001 - 60,000 | 941 (30.9) | 474 (29.5) | 50.3 (45.9, 54.7) | 48.1 (43.7 - 52.6) | **0.84 (0.76, 0.93)** | **0.84 (0.77, 0.93)** |
| 60,001 - 100,000 | 568 (18.7) | 301 (18.8) | 53.0 (48.0, 58.0) | 54.1 (48.9 - 59.2) | 0.89 (0.80, 0.99) | 0.91 (0.82, 1.01) |
| Above 100,000 | 460 (15.1) | 275 (17.1) | 59.8 (54.4, 65.1) | 59.1 (53.3 - 64.6) | Ref | Ref |
| Prefer not to answer | 511 (16.8) | 227 (14.2) | 44.4 (38.8, 50.0) | 45.7 (38.5 - 53.0) | **0.74 (0.66, 0.84)** | **0.80 (0.71, 0.89)** |
| **Employed July 2022** |  |  |  |  |  |  |
| Yes | 1,472 (48.4) | 905 (56.4) | 61.5 (57.9, 65.1) | 57.7 (54.3 - 61.0) | **1.41 (1.32, 1.52)** | **1.31 (1.21, 1.41)** |
| No/DK | 1,570 (51.6) | 699 (43.6) | 44.5 (41.2, 47.7) | 43.5 (39.0 - 48.0) | Ref | Ref |
| **Geographic region** |  |  |  |  |  |  |
| Northeast | 541 (17.8) | 307 (19.1) | 56.6 (51.4, 61.9) | 56.3 (51.4 - 61.2) | Ref | Ref |
| South | 1,153 (37.9) | 603 (37.6) | 52.3 (48.4, 56.2) | 51.5 (47.7 - 55.3) | 0.92 (0.84, 1.01) | 0.94 (0.86, 1.02) |
| Midwest | 630 (20.7) | 323 (20.1) | 51.3 (46.2, 56.5) | 50.7 (45.3 - 56.1) | 0.91 (0.81, 1.01) | 0.95 (0.86, 1.05) |
| West | 718 (23.6) | 371 (23.2) | 51.7 (45.6, 57.8) | 51.0 (45.3 - 56.6) | 0.91 (0.82, 1.01) | 0.96 (0.87, 1.06) |
| **Current vaccination status** |  |  |  |  |  |  |
| Boosted at least once | 1,610 (52.9) | 849 (52.9) | 52.7 (49.4, 56.1) | 53.2 (49.8 - 56.5) | Ref | Ref |
| Fully vaccinated not boosted | 476 (15.6) | 230 (14.3) | 48.3 (42.3, 54.3) | 47.4 (41.0 - 53.9) | 0.92 (0.83, 1.02) | **0.90 (0.81, 0.99)** |
| Not vaccinated | 957 (31.5) | 525 (32.8) | 54.9 (50.1, 59.7) | 53.2 (48.7 - 57.8) | 1.04 (0.97, 1.12) | 0.94 (0.88, 1.01) |
| **Comorbidities** |  |  |  |  |  |  |
| Yes | 1,104 (36.3) | 643 (40.1) | 58.3 (54.2, 62.4) | 59.8 (55.8 - 63.7) | **1.18 (1.10, 1.26)** | **1.27 (1.20, 1.36)** |
| No | 1,938 (63.7) | 961 (59.9) | 49.6 (46.5, 52.6) | 48.4 (45.4 - 51.5) | Ref | Ref |
| **Insured July 2022** |  |  |  |  |  |  |
| Yes | 2,398 (78.8) | 1,277 (79.6) | 53.3 (50.5, 56.0) | 53.7 (51.1 - 56.3) | 1.05 (0.96, 1.14) | **1.21 (1.12, 1.31)** |
| No/DNK | 644 (21.2) | 327 (20.4) | 50.7 (44.8, 56.6) | 46.9 (41.1 - 52.8) | Ref | Ref |
| *Direct standardized for the age and sex groupings in the 2020 US census |  |  |  |  |  |  |
| **Model adjusted for gender and age |  |  |  |  |  |  |
| ***Estimates are adjusted for age only |  |  |  |  |  |  |

In adjusted models, women (versus men), respondents with comorbidities (versus not) and Black NH and API (versus White NH) respondents were less likely to ever have had COVID. Hispanic (versus White NH) respondents and respondents in older age groups (versus 25-34 year olds) were more likely to have ever had COVID. Relative to respondents who were vaccinated and boosted, respondents who were not boosted or not vaccinated were slightly less likely to have ever had SARS-CoV-2 infection. Vaccination status was eliminated from the backward selection model (Supplemental Table 1).

## Discussion

Using a population-representative sample of adults living in the US, we estimate that approximately 18.5 million adults, 7.3% (95% CI: 6.1-8.5%) of the US adult population, was experiencing long COVID (symptoms persisting for more than four weeks after the most recent SARS-CoV-2 infection that were not explained by something else) during the 2-week study period in June-July 2022. Our estimate of 7.3% is consistent with the US Household Pulse Survey ^6^, which estimated 7.5% [7.1-7.9%] of US adults were currently experiencing long COVID. Distressingly, our estimate of prevalence (7.3%) is more than twice that of the UK (2.8%) using the same survey questions.^9^ Of note, the ONS estimates of long COVID are among all UK residents 2 years and older, whereas we estimated long COVID prevalence among US residents 18 years and older. If *no* children or adolescents aged 2-17 years have long COVID in the US, the proportion of all US residents aged 2+ with long COVID would be approximately 5.4% (18 million cases among 331.4 million people living in the United States).^26^ This conservative estimate of the proportion of US residents with active long COVID would remain nearly twice that of the UK (5.4% versus 2.8%). The reasons for this difference are unclear, but could include a lower cumulative incidence of all SARS-CoV-2 infections, including repeat and breakthrough infections, in the UK, e.g. via earlier and higher uptake, as well as better targeting of vaccines and boosters.^27^ Other reasons could be a higher prevalence of risk factors for long COVID (given a SARS-CoV-2 infection) in the US vs. the UK (e.g., a higher prevalence of comorbidities^28^). The causal and predisposing risk factors for long COVID have not yet been fully elucidated. Understanding the reasons for these differences could yield important insights about risk factors, making this an important area for future research.

Long COVID has been associated with a broad range of non-specific symptoms that can overlap with common medical conditions, which makes characterizing the burden of long COVID difficult.^7^ CDC and NIH have established several initiatives to assess the magnitude of long COVID in the US; however, a single surveillance system does not exist, and assessment of long COVID incidence has largely been derived from non-population representative sources.^2,3,6–8^The national public health and medical responses to managing the long term effects of COVID-19 will require understanding long COVID burden, presentation and risk factors. Population-based surveys are an important surveillance tool and supplement to the ongoing national efforts to monitor long COVID.^3^

Identifying groups at higher risk for long COVID and understanding health disparities is essential to managing the long term impacts of the COVID-19 pandemic. Many of the existing studies on long COVID risk factors have been conducted in hospitalized or care-seeking populations, which may overrepresent those with comorbidities. In our study which was not restricted to people accessing medical care, we also observed that female gender and presence of comorbidities were associated with a higher prevalence of long COVID. Age has had an inconsistent association with long COVID in the literature – with some studies finding higher prevalence of long COVID among the oldest age groups.^10,29^ We observed a lower prevalence of long COVID among the oldest (65+ years) age groups, which may be the result SARS-CoV-2 infection risk-reduction behaviors, such as mask wearing of social distancing, higher uptake of vaccines in older US residents, correction of bias from selecting care-engaged populations, and higher rates of COVID mortality in this age group.^8^

Some differences were observed in the risk factors for long COVID versus those for SARS-CoV-2 infection. While female respondents were less likely than male respondents to have ever had SARS-CoV-2 infection, females were almost twice as likely to currently have long COVID. Hispanic respondents were slightly more likely than White NH respondents to have ever had SARS-CoV-2 infection but were substantially less likely to have long COVID. Relative to respondents who were vaccinated and boosted, respondents who were not boosted or not vaccinated were only slightly less likely to have ever had SARS-CoV-2 infection, but they were much more likely to have long COVID. Importantly, we did not capture timing or manufacturer of vaccination relative to SARS-CoV-2 infection or long COVID, and the lack of temporal clarity in these associations require that they be interpreted with caution. For female gender, Hispanic ethnicity and vaccination, the differential risk in long COVID *may not* be due to differences in infection risk and underscores the need for more detailed studies.

While our sample size was small for a detailed examination of those with long COVID, approximately one in three reported their most recent SARS-CoV-2 infection was >12 months prior. We found a significant impact on daily living, which appeared to decrease modestly with the time since the most recent infection. Respondents whose most recent SARS-CoV-2 infection >12 months ago were less likely to report a significant impact on current daily activities relative to respondents who were infected within the past 6 months. While the long-term prognosis and duration of long COVID remain unknown, the natural history of long COVID may be gradual improvement with time for many who have the condition.^8,30^

Our study has other limitations worth noting. First, we measured SARS-CoV-2 infection history over an extended period, which is subject to recall bias. Second, to avoid acute COVID symptoms, we did not assess long COVID among respondents whose most recent SARS-CoV-2 infection was within the past month. However, some of these respondents may have long COVID from an earlier infection, which would result in an underestimation of the current prevalence of long COVID. We also did not measure treatment received for COVID, which may impact prevalence. Third, small sample sizes among population subgroups, including people with long COVID, lowers the precision of estimates across respondent characteristics. Fourth, we are unable to account for non-response bias. If people who have COVID or long COVID were more likely to complete the survey, estimates may be inflated. However, our long COVID prevalence estimates align very well with that of the HPS survey.^6^ Finally, COVID mortality has been shown to be higher among men (versus women), older (versus younger) people, non-Hispanic Black and Hispanic people (versus non-Hispanic White people) and unvaccinated (versus vaccinated) people^31–34^, and estimates of the prevalence of long COVID may be impacted by selection of people who survived COVID, if people who died would have been more likely to go on to develop long COVID (survivorship bias).

Strengths of our study include the representative and probability-based design of the survey, the ability for the survey to reflect outcomes among those who do not access the healthcare system for their COVID infection(s) or long COVID, and the use of similar questions to allow triangulation with other important data sources such as national estimates of long COVID in the UK.

## Conclusions and Public Health Implications

In a national population-representative sample, we observed a very high burden of long COVID and estimated 7.3% of US residents aged 18 years or older (about 18 million adults) had symptoms of long COVID during the two week study period ending July 2, 2022. Our estimate is more than double that of the UK (7.3% versus 2.8%), estimated using a similar methodology, although the UK estimate is for those 2+ years of age.The higher prevalence of long COVID among females relative to males in our study, is not attributable to gender differences in cumulative incidence of prior COVID infections. These differences are important areas of investigation for future research. We found that the duration of long COVID symptoms extends to more than 12 months for many people, but that the impact of long COVID on daily living decreases with time since the most recent infection, suggesting possible improvement in long COVID symptoms over time. Our findings underscore the importance of using a nationally-representative sample to obtain estimates of disease burden when passive surveillance cannot. Such approaches may be helpful for current and future infectious disease outbreaks and pandemics.

## Data Availability

All data produced in the present study are available upon reasonable request to the authors

## Tables and Figures

**Supp Table 1.**
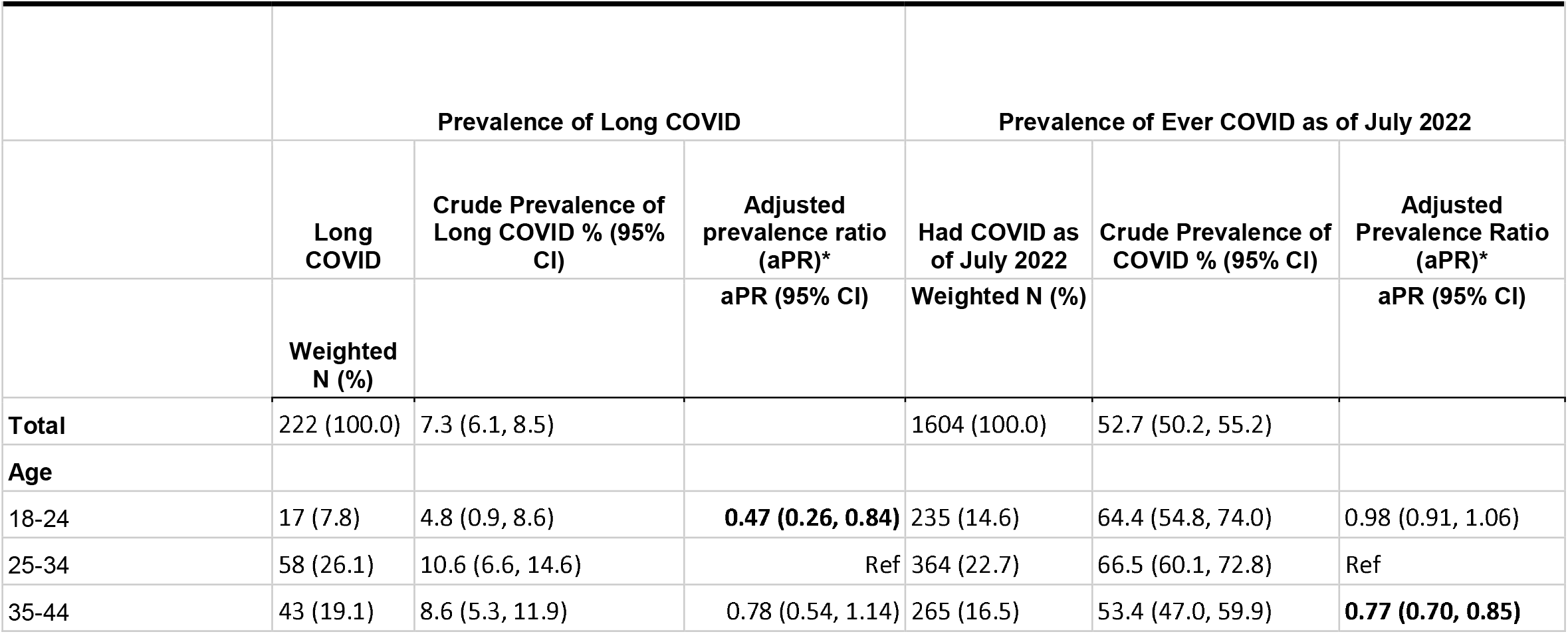
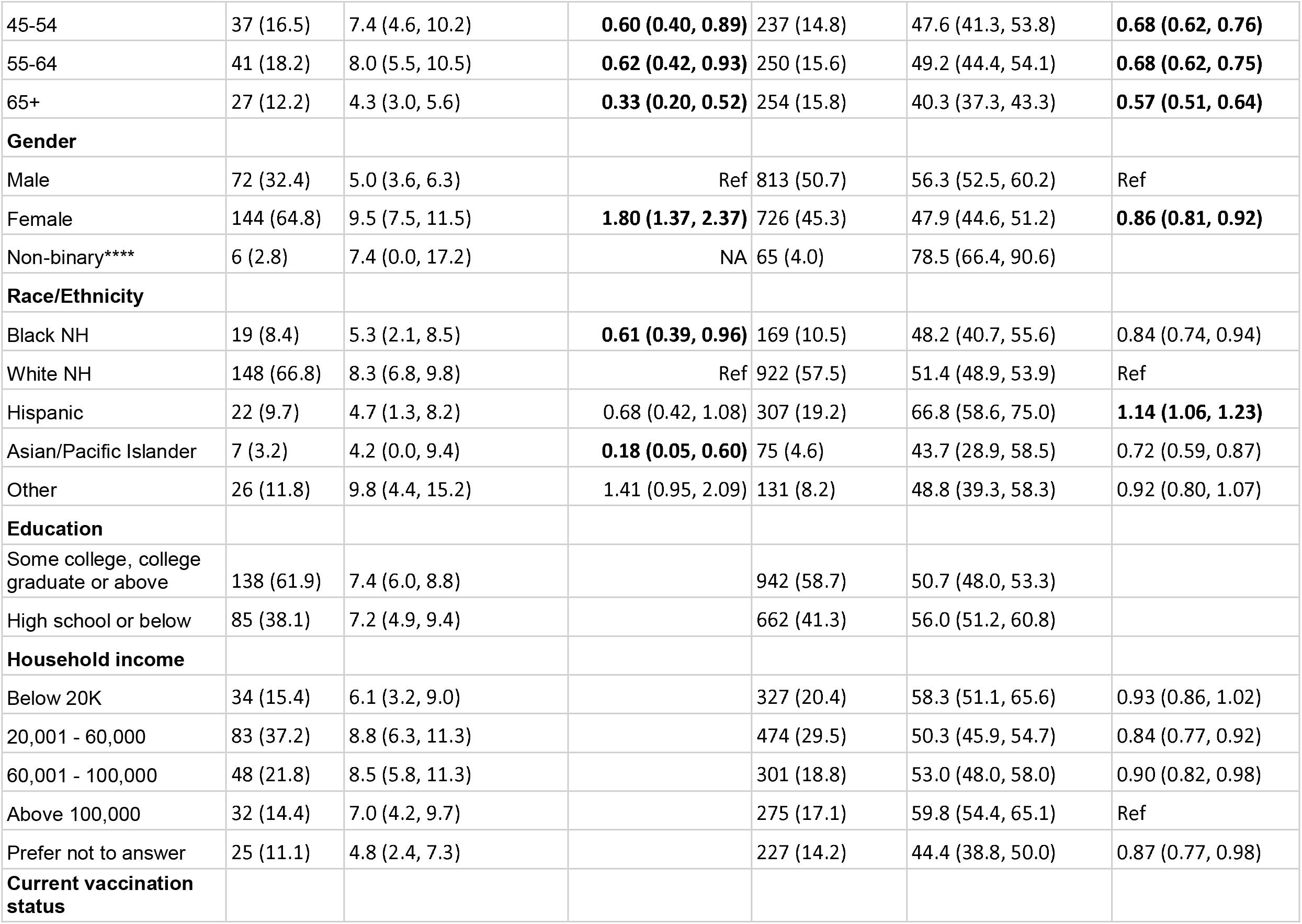
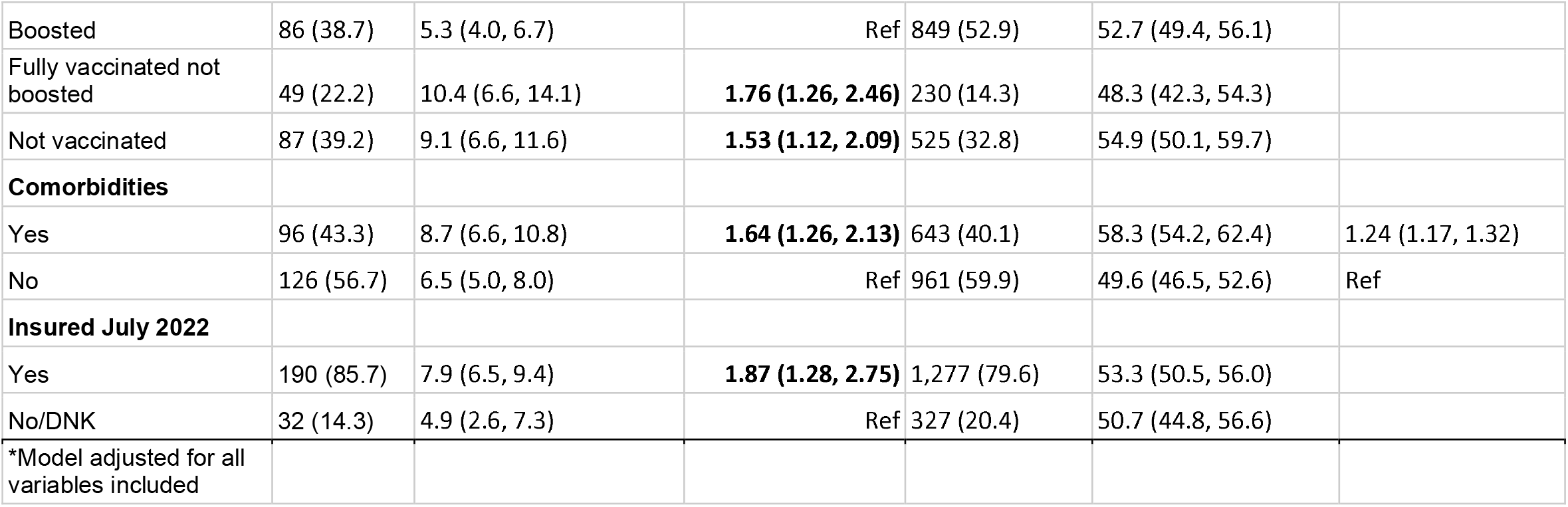
Prevalence and characteristics of US adults with COVID and long COVID, July 2022 - Among Wtd N = 3042 Respondents

|  |  |  |  |  |  |  |
| --- | --- | --- | --- | --- | --- | --- |
|  | Prevalence of Long COVID |  |  | Prevalence of Ever COVID as of July 2022 |  |  |
|  | Long COVID | Crude Prevalence of Long COVID % (95% CI) | Adjusted prevalence ratio (aPR)* | Had COVID as of July 2022 | Crude Prevalence of COVID % (95% CI) | Adjusted Prevalence Ratio (aPR)* |
|  | Weighted N (%) |  | aPR (95% CI) | Weighted N (%) |  | aPR (95% CI) |
| **Total** | 222 (100.0) | 7.3 (6.1, 8.5) |  | 1604 (100.0) | 52.7 (50.2, 55.2) |  |
| **Age** |  |  |  |  |  |  |
| 18-24 | 17 (7.8) | 4.8 (0.9, 8.6) | **0.47 (0.26, 0.84)** | 235 (14.6) | 64.4 (54.8, 74.0) | 0.98 (0.91, 1.06) |
| 25-34 | 58 (26.1) | 10.6 (6.6, 14.6) | Ref | 364 (22.7) | 66.5 (60.1, 72.8) | Ref |
| 35-44 | 43 (19.1) | 8.6 (5.3, 11.9) | 0.78 (0.54, 1.14) | 265 (16.5) | 53.4 (47.0, 59.9) | **0.77 (0.70, 0.85)** |
| 45-54 | 37 (16.5) | 7.4 (4.6, 10.2) | **0.60 (0.40, 0.89)** | 237 (14.8) | 47.6 (41.3, 53.8) | **0.68 (0.62, 0.76)** |
| 55-64 | 41 (18.2) | 8.0 (5.5, 10.5) | **0.62 (0.42, 0.93)** | 250 (15.6) | 49.2 (44.4, 54.1) | **0.68 (0.62, 0.75)** |
| 65+ | 27 (12.2) | 4.3 (3.0, 5.6) | **0.33 (0.20, 0.52)** | 254 (15.8) | 40.3 (37.3, 43.3) | **0.57 (0.51, 0.64)** |
| **Gender** |  |  |  |  |  |  |
| Male | 72 (32.4) | 5.0 (3.6, 6.3) | Ref | 813 (50.7) | 56.3 (52.5, 60.2) | Ref |
| Female | 144 (64.8) | 9.5 (7.5, 11.5) | **1.80 (1.37, 2.37)** | 726 (45.3) | 47.9 (44.6, 51.2) | **0.86 (0.81, 0.92)** |
| Non-binary**** | 6 (2.8) | 7.4 (0.0, 17.2) | NA | 65 (4.0) | 78.5 (66.4, 90.6) |  |
| **Race/Ethnicity** |  |  |  |  |  |  |
| Black NH | 19 (8.4) | 5.3 (2.1, 8.5) | **0.61 (0.39, 0.96)** | 169 (10.5) | 48.2 (40.7, 55.6) | 0.84 (0.74, 0.94) |
| White NH | 148 (66.8) | 8.3 (6.8, 9.8) | Ref | 922 (57.5) | 51.4 (48.9, 53.9) | Ref |
| Hispanic | 22 (9.7) | 4.7 (1.3, 8.2) | 0.68 (0.42, 1.08) | 307 (19.2) | 66.8 (58.6, 75.0) | **1.14 (1.06, 1.23)** |
| Asian/Pacific Islander | 7 (3.2) | 4.2 (0.0, 9.4) | **0.18 (0.05, 0.60)** | 75 (4.6) | 43.7 (28.9, 58.5) | 0.72 (0.59, 0.87) |
| Other | 26 (11.8) | 9.8 (4.4, 15.2) | 1.41 (0.95, 2.09) | 131 (8.2) | 48.8 (39.3, 58.3) | 0.92 (0.80, 1.07) |
| **Education** |  |  |  |  |  |  |
| Some college, college graduate or above | 138 (61.9) | 7.4 (6.0, 8.8) |  | 942 (58.7) | 50.7 (48.0, 53.3) |  |
| High school or below | 85 (38.1) | 7.2 (4.9, 9.4) |  | 662 (41.3) | 56.0 (51.2, 60.8) |  |
| **Household income** |  |  |  |  |  |  |
| Below 20K | 34 (15.4) | 6.1 (3.2, 9.0) |  | 327 (20.4) | 58.3 (51.1, 65.6) | 0.93 (0.86, 1.02) |
| 20,001 - 60,000 | 83 (37.2) | 8.8 (6.3, 11.3) |  | 474 (29.5) | 50.3 (45.9, 54.7) | 0.84 (0.77, 0.92) |
| 60,001 - 100,000 | 48 (21.8) | 8.5 (5.8, 11.3) |  | 301 (18.8) | 53.0 (48.0, 58.0) | 0.90 (0.82, 0.98) |
| Above 100,000 | 32 (14.4) | 7.0 (4.2, 9.7) |  | 275 (17.1) | 59.8 (54.4, 65.1) | Ref |
| Prefer not to answer | 25 (11.1) | 4.8 (2.4, 7.3) |  | 227 (14.2) | 44.4 (38.8, 50.0) | 0.87 (0.77, 0.98) |
| **Current vaccination status** |  |  |  |  |  |  |
| Boosted | 86 (38.7) | 5.3 (4.0, 6.7) | Ref | 849 (52.9) | 52.7 (49.4, 56.1) |  |
| Fully vaccinated not boosted | 49 (22.2) | 10.4 (6.6, 14.1) | **1.76 (1.26, 2.46)** | 230 (14.3) | 48.3 (42.3, 54.3) |  |
| Not vaccinated | 87 (39.2) | 9.1 (6.6, 11.6) | **1.53 (1.12, 2.09)** | 525 (32.8) | 54.9 (50.1, 59.7) |  |
| **Comorbidities** |  |  |  |  |  |  |
| Yes | 96 (43.3) | 8.7 (6.6, 10.8) | **1.64 (1.26, 2.13)** | 643 (40.1) | 58.3 (54.2, 62.4) | 1.24 (1.17, 1.32) |
| No | 126 (56.7) | 6.5 (5.0, 8.0) | Ref | 961 (59.9) | 49.6 (46.5, 52.6) | Ref |
| **Insured July 2022** |  |  |  |  |  |  |
| Yes | 190 (85.7) | 7.9 (6.5, 9.4) | **1.87 (1.28, 2.75)** | 1,277 (79.6) | 53.3 (50.5, 56.0) |  |
| No/DNK | 32 (14.3) | 4.9 (2.6, 7.3) | Ref | 327 (20.4) | 50.7 (44.8, 56.6) |  |
| *Model adjusted for all variables included |  |  |  |  |  |  |

## Funding

Funding for this project was provided by the CUNY Institute for Implementation Science in Population Health (cunyisph.org).

## Acknowledgements

The authors wish to acknowledge the survey participants and Consensus Strategies for completing survey sampling and data collection.

## Appendix 1 (Survey design)

### Sampling Frame

A sampling frame of 254,297,978 Residents of the United States consisting of 105,469,157 mobile numbers with an additional 60,126,857 landlines. Two stratified proportionate randomized population-based samples were drawn for this study, n=90,000 mobile numbers and n=50,000 landlines. An National opt-in Online Panel provided by Consensus Strategies was used in the study. A total sample of n=3,042 was utilized with a +/- 3 percent margin of error. Data was collected June 30 - July 2, 2022.

### Multi-mode data collection design

Short message service (SMS) aka text messages were sent using SMS platform. The respondents were sent a personalized first name text message which included a link to the survey and an opt-out option. The respondents had the option to reply to the SMS text with any queries. Data was verified by IP address and scrubbed against the original survey sample.

Interactive voice response (IVR) aka robo-poll messages were sent to landlines. The respondents were able to answer the survey questions using the touch tone keypad on their phones.

The opt-in online panel was created by Consensus Strategies and participants were paid an incentive to complete the surveys of up to $2. Respondents were verified by payment information.

### Survey weighting

The survey was weighted using an iterative weighting method (raking) to marginal proportions of race, ethnicity, age, self-identified sex, and education by U.S. region. The samples (landline, online, mobile) were normalized at the region level based on sex, age, gender, education, race and sample size then combined and weighted back based on the proportion of the region to the overall population and the other demographics. The sum of the weights equals the sample population (n=3,042).

Demographic weights were created based on the American Community Survey 5-year estimates and <u>2020 US Census</u>. The inference population is 254,297,978 million adult residents.

### Response rates

Our overall combined response rates across all modalities was 7.2%. The response rate was 6.2% for random digit dial to landline, 0.9% for cell phone, and 86.5% for opt-in online panel. The response rate reflects the proportion of complete respondents among eligible participants in the sampling frame. For context, we also included response rates for the Household Pulse Survey (HPS), an online survey sampling households from the Census Master Address File with an email or cell phone number. While our response rates are comparable, the HPS methodology calculates the response rate based on completes and sufficient partial interviews, compared to our rates which are based on complete interviews (i.e., more conservative rates)

|  | ***National COVID-19***<br>***Survey (June-July 2022)*** | ***HPS***<br>***202*** | ***HP***<br>***S*** | ***HP***<br>***S*** | ***HP***<br>***S*** | ***HP***<br>***S*** |
| --- | --- | --- | --- | --- | --- | --- |

| *Mod*<br>*e* | *Landli*<br>*ne* | *C*<br>*ell* | *Onli*<br>*ne*<br>*opt-*<br>*in*<br>*pane*<br>*l* | *Combi*<br>*ned* | *2*<br>*Jun*<br>*29-*<br>*Jul*<br>*11* | *202*<br>*2*<br>*Jun*<br>*1-*<br>*Jun*<br>*13* | *20*<br>*20*<br>*We*<br>*ek*<br>*1* | *202*<br>*0*<br>*We*<br>*ek*<br>*2* | *202*<br>*0*<br>*We*<br>*ek*<br>*3* |
| --- | --- | --- | --- | --- | --- | --- | --- | --- | --- |
| *RR* | 6.2 | 0.<br>9 | 86.5 | 7.2 | 5.7 | 6.2 | 3.8 | 1.3 | 2.3 |

## Appendix 2 (Survey questionnaire)

### Survey on recent COVID exposure, COVID infection, and testing behaviors in the United States

Hello, this is XYZ with a brief public policy survey. At no time will we try to sell you anything. We are just interested in your opinions, and you can drop out at any time.

To begin, what language would you like to take this survey in?

1. English
2. Español

#### The following questions will ask about COVID exposure in the past 2 weeks

1. In the past 2 weeks, have you experienced any COVID-like symptoms (e.g., 100 degrees fever or higher, chills, cough, sore throat, fatigue, headache, shortness of breath, congestion or runny nose, muscle aches, loss of smell or taste, nausea, or diarrhea)?
  a. Yes
  b. No
  c. Don’t know/not sure
2. In the past 2 weeks, were you aware of an exposure you had to someone who had COVID-like symptoms or tested positive for COVID-19?
  a. Yes
  b. No
  c. Don’t know/not sure

#### The following questions will ask about COVID testing in the past 2 weeks

3. In the past 2 weeks, have you taken an at-home rapid test for COVID-19? (a rapid at-home test allows you to collect your own sample and get results within minutes at home)
  a. Yes, Tested Positive
  b. Yes, Tested Negative
  c. No, I have not tested
4. In the past 2 weeks, have you taken a rapid antigen or PCR test for COVID-19 from a healthcare or testing provider?
  a. Yes, Tested Positive
  b. Yes, Tested Negative
  c. No, I have not tested [skip to 6]
5. *If tested with a healthcare provider*. When you tested with a healthcare or testing provider, which type of test did you receive?
  a. Rapid test/point of care test
  b. PCR test
  c. Both
  d. Not sure/don’t know
6. Prior to June 15th 2022, did you ever have COVID-19 infection, either diagnosed by a healthcare or testing provider, or based on a positive at-home rapid test?
  a. Yes, once
  b. Yes, more than once
  c. No, but I am pretty sure that I had COVID
  d. No, I don’t think I have ever had COVID [skip to 8]
  e. Don’t know/not sure [skip to 8]
7. When was the last time you had COVID?
  a. Within the last month (if you currently have COVID, choose this option)
  b. 1-3 months ago
  c. 3-6 months ago
  d. 6-12 months ago
  e. >12 months ago
  f. Don’t know/not sure

#### Long COVID

8. Would you describe yourself as having ‘long COVID’, that is you experienced symptoms such as fatigue, difficulty concentrating, shortness of breath more than 4 weeks after you first had COVID-19 that are not explained by something else?
  a. Yes
  b. No [skip to 10]
  c. Don’t know/not sure [skip to 10]
9. Does this reduce your ability to carry-out day-to-day activities compared with the time before you had COVID-19?
  a. Yes, a lot
  b. Yes, a little
  c. Not at all
  d. Don’t know/not sure

#### Respondent Characteristics

10. Do you currently have any kind of health care coverage, including health insurance, prepaid plans such as HMOs, or government plans such as Medicaid or Medicare, or Indian Health Service?
  a. Yes
  b. No
  c. Don’t know/not sure
11. Do you have any of the following conditions that could increase the severity of COVID-19: cancer, diabetes, obesity, COPD or lung disease, liver disease, heart disease, high blood pressure, a recent organ transplant, or an immunodeficiency)?
  a. Yes
  b. No
  c. Don’t know/not sure
12. Have you been fully vaccinated against COVID-19? [Either 2 doses of mRNA vaccine series (Moderna or Pfizer) or a single dose of Johnson and Johnson COVID-19 vaccine]
  a. Yes
  b. No (go to 14)
  c. Don’t know/not sure (go to 14)
13. If you have been fully vaccinated, have you also received at least one coronavirus booster?
  a. Yes, more than 5 months ago
  b. Yes, within the past 5 months
  c. No
14. If not fully vaccinated OR not boosted: Do you plan to get a vaccine dose or booster in the next two weeks?
  a. Yes
  b. No
  c. Don’t know/not sure
15. What is your age?
  a. 18-24
  b. 25-34
  c. 35-44
  d. 45-49
  e. 50-54
  f. 55-64
  g. 65-74
  h. 75 +
16. How do you currently identify your gender? Do you identify as …
  a. Male
  b. Female
  c. Gender non-binary
  d. other
17. Which one of the following would you use to describe yourself ?
  a. Latino/a, or of Hispanic or Spanish origin
  b. White
  c. Black or African American
  d. Asian, Native Hawaiian or Other Pacific Islander
  e. American Indian/Alaska Native
  f. More than one race
  g. other
18. What is the highest grade or year of school you completed?
  a. Less than high school
  b. Grade 12 or GED (High school graduate)
  c. College 1 year to 3 years (Some college or technical school, associate degree)
  d. College 4 years or more (College graduate)
19. Are you currently employed for wages or salary?
  a. Yes
  b. No
  c. Don’t know/not sure
20. What is your household’s annual income?
  a. $20,000 or less
  b. Between $20,001 - $40,000
  c. Between $40,001 - $60,000
  d. Between $60,001 - $80,000
  e. Between $80,001 - $100,000
  f. Above $100,000
  g. Prefer not to answer
  h. Don’t know/not sure

## Notes

### Competing Interest Statement

MMR, DN and SGK receive support from Pfizer

